# COVID-19 RISK EVALUATION AND TESTING STRATEGIES BASED ON CONTACT TRACING NETWORK AND INFORMATION ANALYSIS

**DOI:** 10.1101/2020.11.30.20240762

**Authors:** Ankush Kumar

**Affiliations:** Univ. Lille, CNRS, Centrale Lille, Univ. Polytechnique Hauts-de-France, UMR 8520 - IEMN - Institut d’Électronique de Microélectronique et de Nanotechnologie, F-59000 Lille, France

**Keywords:** COVID-19 risk evaluation, COVID-19 testing, Contact tracing

## Abstract

Contact tracing and efficient testing can have an imperative part in mitigating the COVID-19 spread, with minimal social and economic disruption. Testing serves many purposes: isolating the COVID-19 positive tested individuals, identifying the contacts at the risk, and locating the hotspots and safe zones for administrative planning. However, it is a challenging task to identify the right individuals for the test in view of the high COVID -19 spread, a large number of presymptomatic and asymptomatic cases, and limited testing capabilities. The individuals for COVID -19 are currently identified based on direct-contact, travel history, and symptoms, which are more individualized and do not explicitly include a group risk assessment, and in turn, do not preclude the transmission from the superspreaders. Policymakers need to limit testing in the shortage of test resources, and focus on gaining the most information from the tests performed. In this work, we introduce a protocol for the identification of the group of individuals to be tested for acquiring maximum risk information of a community with minimum individual tests performed. Firstly, an algorithm is proposed to determine the risk profile of all the individuals in the community by incorporating serial and parallel pathways of the infection transmission considering multiple steps of transmission. Next, we consider several potential groups that could be tested from the community, and analyze them one by one for their comparison. In a group, few individuals can be positive, and the remaining few can be negative, generating sets of several test-outcomes with unequal probabilities. The protocol involves the probability calculation and reassessment of the network’s risk profile in all the test output cases. Finally, the best group is identified in all the groups studied, in which risk profiles between post and pre-test are maximally different. The analysis shows that in general, information increases with an increase in the group size. Notably, a strategically chosen small group may provide more information from the test results, than a standard larger group. The proposed systematic strategy would help in the selection of the right individuals for the testing, and in extracting far more information from the minimum samples, to effectively aid the epidemic mitigation. The protocol is generic, and can also be applied to any other epidemic spread in the future.

## 1 Introduction

COVID-19 is an unprecedented challenge for mankind; affecting health care, the global economy, movement, and social life. Various stringent policies have been placed to mitigate the spread at the cost of shutting business and social events, which means that these initiatives are not permanent and can only be enforced in risky zones. [1] In contrast to all other past epidemics, such as SARS and MERS, asymptomatic cases of COVID-19 in the population are very high, and it is necessary to rely only on specialized testings. [2] In the present situation, contact tracing and well-planned testing are the most important public health interventions available, also recommended by World Health Organization. [3, 4, 5] Various apps are launched by several countries, such as Trace Together by the Singapore govt, Aarogya Setu by the Indian govt, TousAntiCovid by French govt, etc. which are based on Bluetooth based entries to capture the possible interactions, for acquiring the information of interaction with an infected individual. [6] As an example, the TousAntiCovid alerts the individuals who have been in close contact with a reported COVID-19 user for exposure at less than one meter, lasting 15 minutes or more. Most importantly, the contact tracing and testing helps in understanding the statistics of community spread, required for policymakers to contain it. [7, 8, 9, 10] And if vaccine or cure for the COVID-19 is possible in the near future, there is always a need to focus on contact tracing to target individuals for vaccination or meditation. Various testing strategies have been discussed for effectiveness, such as bidirectional contact tracing [11], pool testing [12, 13], and testing at the exit of the quarantine period than the entry. [14] Effectiveness of contact tracing depends on the number of contaminated individuals, testing quickness, fraction, and rapidness in contact tracing and quarantine, and tracking the further transmission. [15, 16, 10] The individuals for COVID -19 are primarily identified based on direct-contacts, travel history, symptoms, and random sampling. Mass scale testing in most of the countries is impractical to conduct due to higher testing costs, the flow of new entrants into the community [17], and the ability of the person tested to become contaminated in the future. The infection spread is largely due to superspreaders in the community who transmit the infection to several individuals, [18] these cases can only be prevented by detailed network analysis. [19] Thus, it is very important to post-process the contact tracing data, and make strategies for testing and isolating individuals at the risk. Policymakers should have strategies to acquire maximum spread information using minimum tests conducted in the community. However, the detailed risk evaluation based on a network level with several multi serial and parallel connections is still an ongoing challenge. Currently, the goal of a test is largely on an individual level, and to obtain the positivity rate of the community, but does not provide a detailed risk assessment of the community. Apps such as Aarogya Setu inform the users of the exposure to COVID-19, by testing their proximity to known patients, [20] while considering only single step transmission. As per our knowledge, only Guttal et al. [21] have looked at the risk determination of the contacts, however, it uses the Gillespie simulations, which may not be easy to perform and does not consider the weighted connections. The present work introduces the strategies to evaluate the risk profile of the community, identify individuals for tests, and propose testing protocols in the direction of acquiring the maximum information with a minimum number of tests.

## 2 Results and Discussions

Consider a social network with individuals represented as nodes, and edges representing the transmission probability *T*_*i*_ → _*j*_ of infection between the individuals. Figure 1a represents a typical network with 13 individuals, represented as alphabets from A-K, with red and blue nodes representing the infected and the susceptible individuals respectively. The corresponding connections values (0-1), represent the probability of disease transmission between the individuals. The transfer probability can be obtained from app-based data or manual contact tracing questionnaire considering the time of mutual interaction between the individual, possibility of spread, and decaying the weight over time. In this work, we do not focus on the network construction based on contact tracing, but rather on post-analysis of the network, which would be important for the risk assessment and suggested testing strategies. For the first step of transmission, 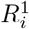 shown in Figure 1b, nodes immediately linked to the infected nodes may be at risk of infection, the risk value proportional to the total of transmission strengths from neighboring infected nodes. The risk value (0-1) is the risk that the transmission from the reported cases in the community could potentially infect a person. In this manuscript, the risk value is based only on an overview of the contact tracing network taking into account the network’s direct and indirect transmission route. As an example, there are 2 infected individuals connected to node B, i.e. node J and node M, with interaction weights of 0.2 and 0.26 respectively, thus the probability of infection of node K is 0.46. Figure 1(b) displays all individuals’ risk profiles, calculated similarly, with the node values and color intensity as the risk. Nodes with higher risk values can be potentially infected nodes, and in turn, may pass the infection to their respective neighbors in the next step. The risk of *i* individual for *s* step transmission 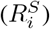 considering all neighbors *j* with their corresponding risk values 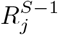 at *S* − 1 steps can be calculated as,

**Figure 1:**
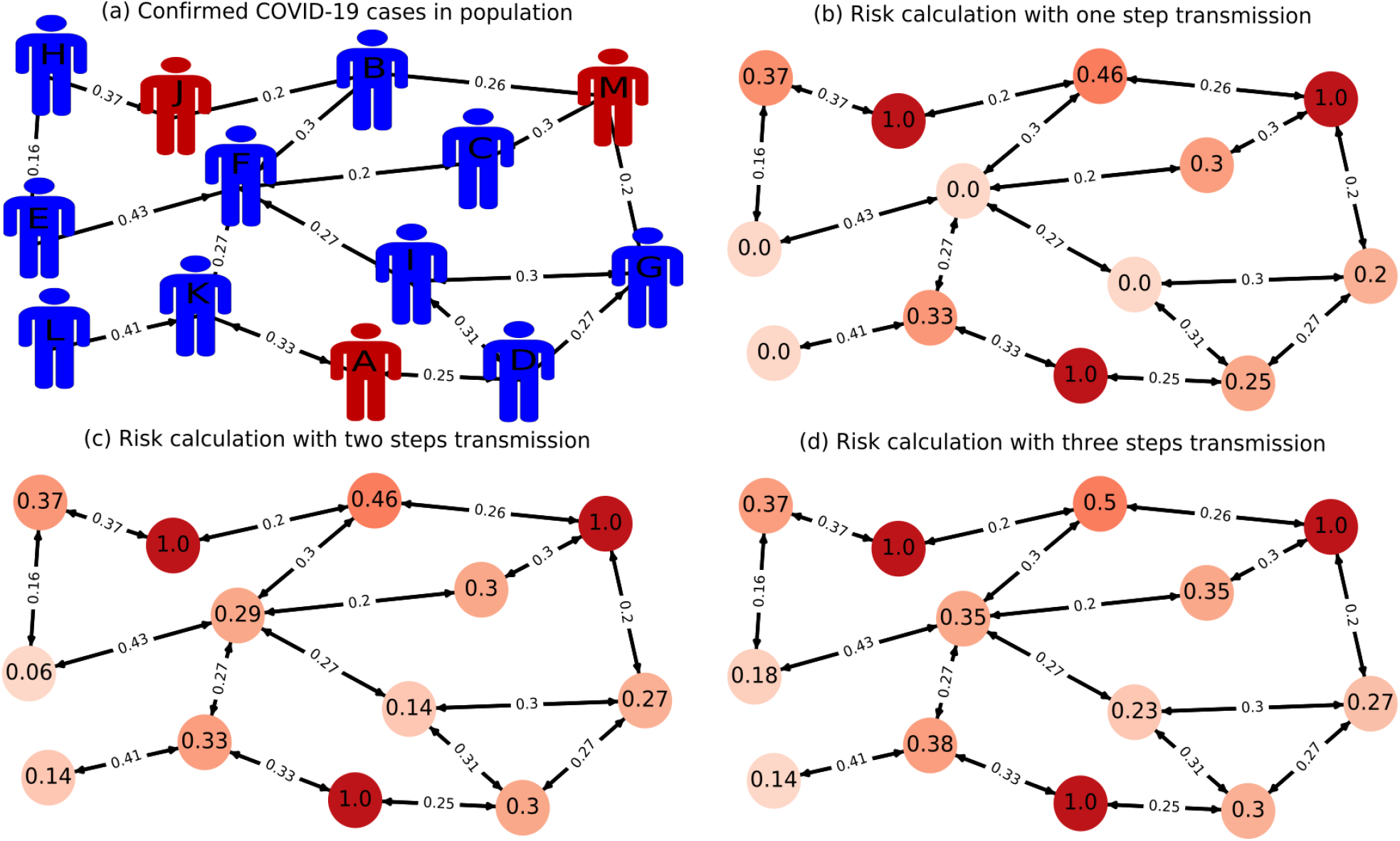
(a) Contact tracing network of a community, with red nodes representing the infected individuals and blue nodes displaying the susceptible population. The legends on nodes are the individual identities represented as alphabets A-K, the values on the edges represent the transmission probability of the infection between the nodes. The network is studied in the manuscript as an example network. (b) Risk profile evaluation based on a single step transmission with risk values displayed as color intensity, and label values from 0 to 1 representing the probability of infection. (c-d) Risk profile with two steps and three steps transmission. The risk values are obtained using algorithm 1 proposed in the manuscript.

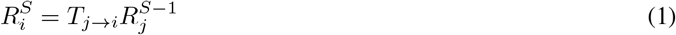

Figure 1c indicates the risk profile with the two steps transmission; it raises the risk values, adding several people without risk (see nodes E, F, I, L). As an example, the node G is connected to D, I and M with transmission values, *T*_*D*→*G*_ as 0.27, *T*_*I*→*G*_ as 0.3 and *T*_*M*→*G*_ as 0.2 and risk values from the first step (*s* = 1) (Figure 1b) 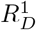 as 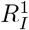 as 0 and 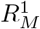 as 0.25, hence the integrated risk at G, 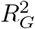 based on Eq. 1 can be written as 0.27. The study can be useful for finding and alerting persons with a risk value exceeding a certain level; individuals with a risk threshold exceeding say 0.3 can be alerted which are B (0.46), H (0.37), K (0.33), C (0.3), and D (0.3). Note that a variable threshold value dependent on age and medical history may also be incorporated in the realistic case. One can easily make a histogram of risk values to quantify the spread in the community. Furthermore, the simulations can be re-run for long-term forecasts of several transmission phases as seen in Figure 1d for three transmission steps demonstrating maximum risk at the nodes: B, K, H, C and F. Algorithm 1 shows the detailed steps of the computation, the parallel connections terminating at a node are added, while the serial connections are multiplied, constraining the number of steps, and total risk value not exceeding unity. Note that, the algorithm, avoids self-loops in accordance with the physical situation, that is, infection by *i* to *j* and back *j* to *i* (by any mode) is not allowed. The time scales of transmission of epedemic can be integrated in order to predict the time evolution of infected population. Although the code can anticipate multi-step predictions, we have confined the transmission to only two stages in further studies, for the simplicity of the readers.

It is important to test the potentially infected individuals for controlling the epidemic spread. Nevertheless, in a shortage of test resources, policymakers need to minimize the tests and focus on acquiring maximum information from the tests. Herein, we try to identify the group of individuals, who should be tested for the maximum information. For a simple illustration, we pick three groups with two individuals in each: [C, E], [G, H], and [D, F] for testing, and compare their analysis. For each group, there are 4 possible outcome cases: both of the individuals are tested positive, none is positive or either one of them is positive, with *P*_*c*_ being the probability of a particular output case. The probability of an individual to be positive depends on its risk value. As an example, for the testing of [C, E] the probability of the positive outcome of C is 0.3,(Figure 1c) and E being positive is 0.06 (Figure 1c), thus the probability of C and E both being positive is the product of probability i.e. 0.018. The individual probability for C and E to be negative is 0.7 (1-0.3) and 0.94 (1-0.06), which is 0.658, shown in Figure 2 in two decimal values. Thus, the probability of an outcome case (*P*_*c*_) can be written in terms of individual risk values of tested positive and negative individuals as:

**Figure 2:**
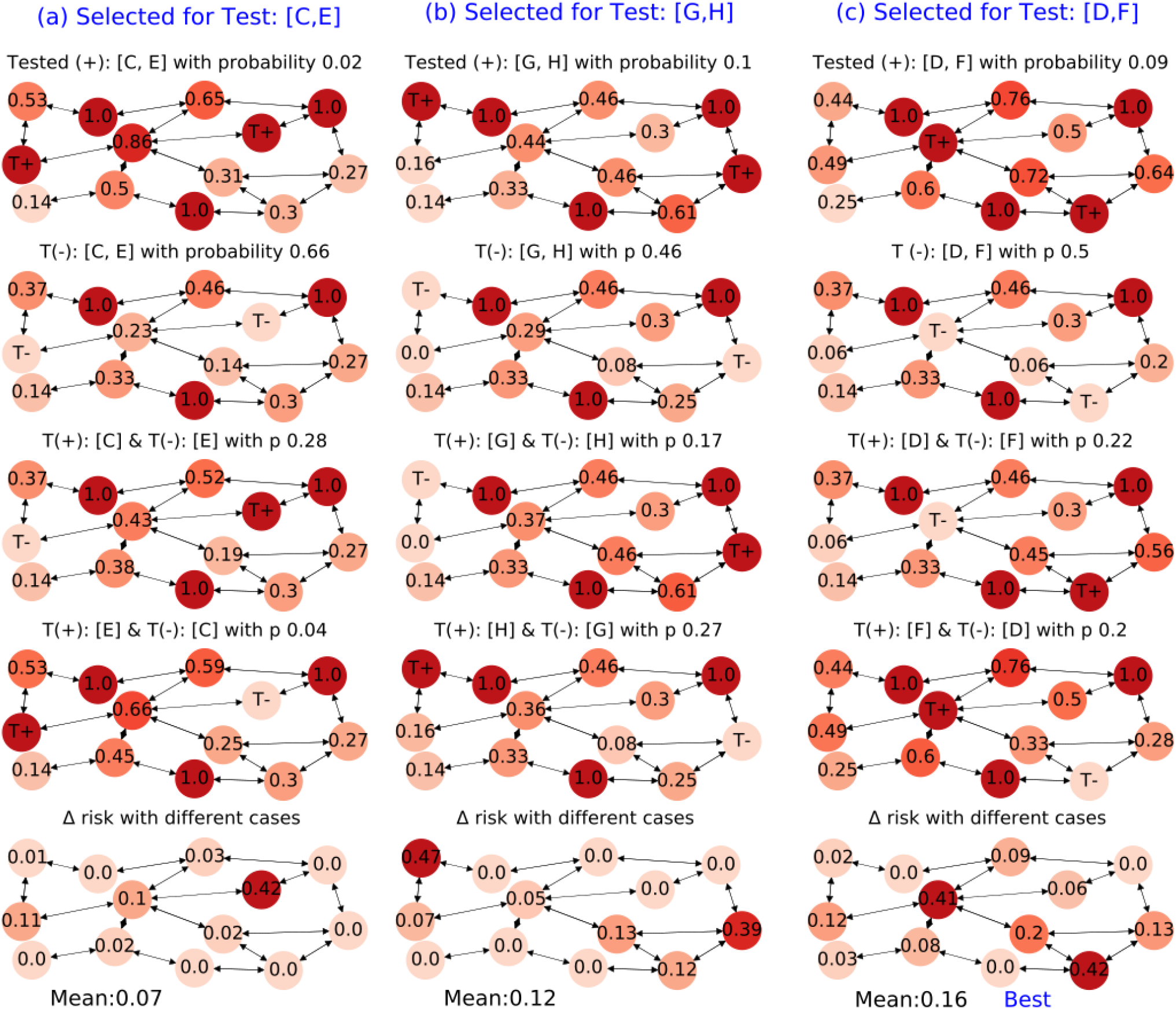
Comparision of testing strategy of three group-sets: (a) [C,E], (b) [G,H] and (c) [D,F] for the population shown in Figure 1a. All the cases of test outcomes are shown in the same column: (i) both being positive, (ii) both being negative (iii) the first individual being positive and the second being negative, and (iv) the second individual being positive and the first being negative. The probability of a positive test individual outcome is based on its risk while the probability of a negative outcome is 1-Risk value, computed in Figure 1b. The individual probabilities of the test outcomes are multiplied to obtain group test probability. The tested positive individual is represented by T+ and tested negative is represented by T-. The risk profile for all these cases is re-calculated and represented as node values. The risk variation for all group tests is compared, considering all the above cases with corresponding probabilities of occurrence. The best group is found to be [D, F], having maximum risk variation with the tests, which implies maximum information with the tests.

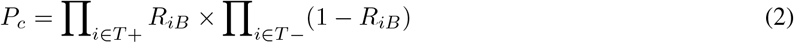

The risk value of the tested positive individual is updated as 1, and for the negative tested individual as zero. Since all the cases are having unequal probabilities (*p*_*c*_) and risk values (*R*_*iC*_), the risk variation of a node, Δ*R*_*i*_ can be written as

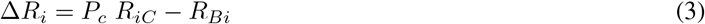

### Algorithm 1: Algorithm to calculate risk based on confirmed cases

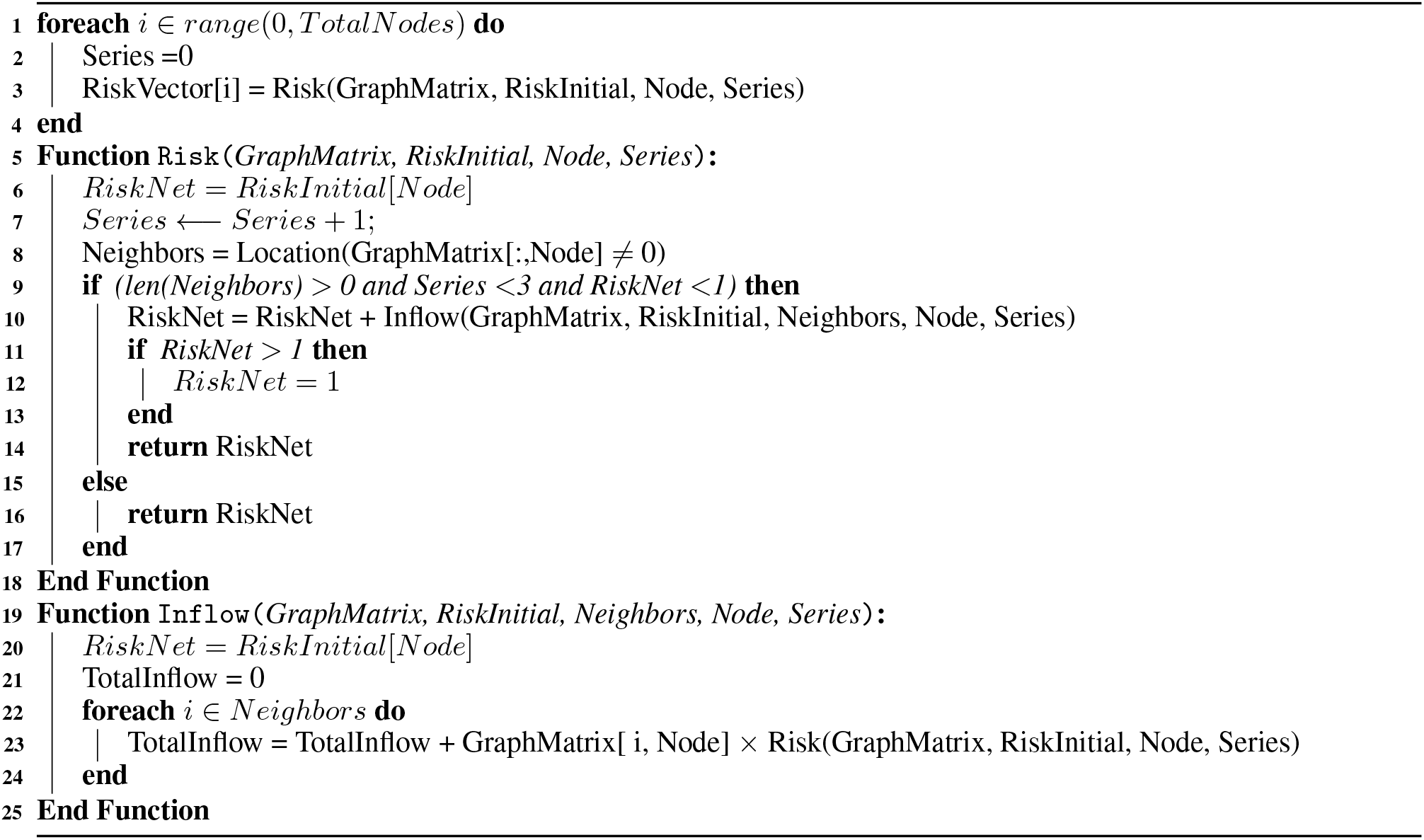

As an example, for both [C, E] to be positive, the risk value (*R*_*B*_1) of B node is 0.65 (with *p*_1_ = 0.02), for both being negative (*R*_*B*2_), the value is 0.46 (with *p*_2_ = 0.66) and positive test of C alone *R*_*B*3_ is 0.52 (with *p*_3_ = 0.28) and for E alone (*R*_*B*4_) is 0.59 (with *p*_4_ = 0.04). Thus, the test outcome, significantly changes the risk profile in different ways as compared to before the test (*R*_*B*0_), and Δ*R*_*B*_ is 0.03. The risk profiles, for different test outputs can help in alerting differernt individuls. The varaiation in risk values, is very sensitive to the selection of individuls. Δ*R*_*B*_ values for [C,E], [G,H] and [D,F] groups are 0.03, 0 and 0.09, higher the value higher the importance of a test. The mean variation in the risk can be written in the terms of sussaptible popuation (S):

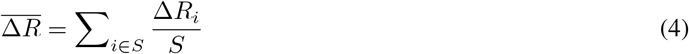

The objective of a test in general is to acquire information about a system, which the observer does not know before carrying out the test. In this particular case, if the risk profile variation with a test is negligible, it means there is no benefit of a test. As an example, when testing the (a) group [C, E], Δ*R* values in the risk profile map of the network is negligible, with significant variation (>0.1) in only a few nodes: C (0.42), E (0.11) and F(0.1), implying not much network information is obtained by testing the group [C, E]. While in the case of testing the (c) group [D, F], the risk variation in many nodes: D(0.42), F (0.41), I (0.2), E(0.12), and G(0.13) is significant, suggesting the possibility of more information by testing the group [D,F]. The mean Δ*R* values for [C,E], [G,H] and [D,F] are 0.07, 0.12 and 0.16, the higher value of mean risk variation representing more global information obtained from the test. The different test outcomes can help in alerting the right individuals based on an updated risk profile. As an example, while testing the group [D, F], if the test output is case 3 (Tested positive:D and Tested negative: F) having a probability of 0.22 to occur, one can alert G (0.56), B (0.46), I (0.45), H (0.37), and K (0.33). While, if the test output is case 4 (Tested positive: F and Tested negative:D) having a probability of 0.2 to occur, the individuals B(0.76), K (0.6), C (0.5), E (0.49), H (0.44), and I (0.33)) should be alerted. It is clear the risk profile in both these cases are significantly different, demonstrating high information provided by the test output. Thus, it is advisable to select [D, F] for the test, while [C, E] provides the minimum information. Algorithm 2 shows the detailed steps of the systemic selection of individuals for the proposed testing strategy. Notice the selection of [D, F] is not easily intuitive, showing the significance of the algorithm introduced, which takes into account different aspects of network transmission.

### Algorithm 2: Algorithm to select individuals for testing

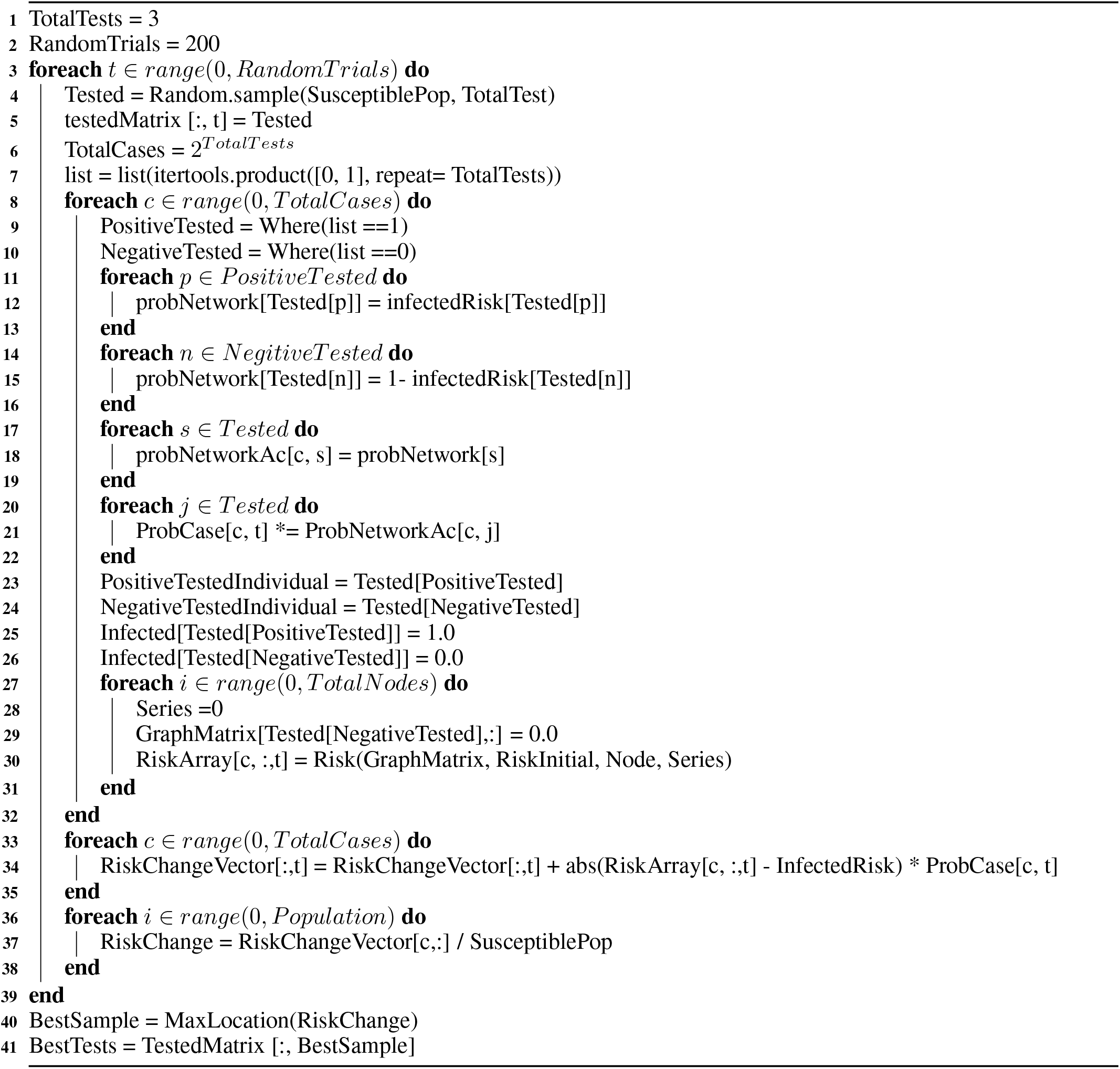

Figure 3 shows examples of selected test groups with a variable number of individuals in each group. The total number of output cases with *N* individuals in the group is 2^*N*^. It is found that the risk variation for a group of 4,3, 2 and 1 individuals are maximum for [B, D, H, K] (0.26), [B, D, K] (0.21),[D, F] (0.16)and [E] (0.09), representing information increases with the increase in the group size. However, note that the risk variation depends strongly on the selection set of the group. As an example, the risk variation of a 2 individuals group [D, F] is higher than 3 individuals [G, H, I] and 4 individuals group [C, E, F, I]. A systematically selected smaller group can offer more information, than an unplanned selected bigger group. As the computation considers all the attributes of the network, thus one need not rely on betweenness, degree, or any such network-feature for selecting the nodes. The protocol is automated using the proposed algorithm and thus can be used at different institutional levels, with minimal training of the policy-user. Though we have used COVID-19 as a specific challenge to handle, nevertheless, the approach is generic and can be applied to any similar epidemic, without any specific modification.

**Figure 3:**
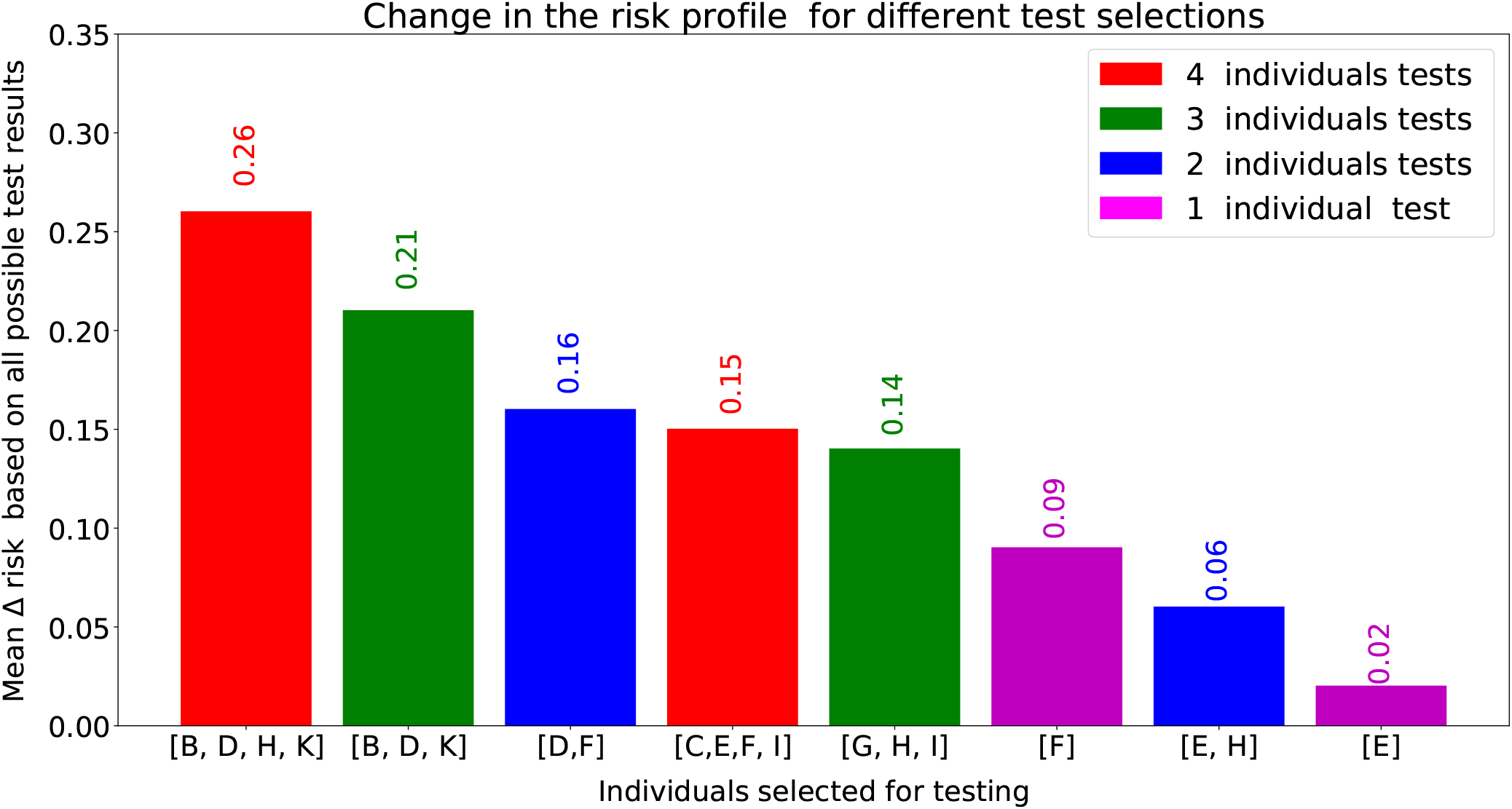
Comparision of change in risk values by considering all possible test outcomes, condidering their probability. Higher the value of risk change, more is the information from the test. Thus the selected group is apprpriate for the tesing.

## 3 Conclusion

In the manuscript, we have developed a model for the evaluation of the epidemic risk profile in the community and discussed effective testing strategies for transmission chain mitigation. A simple model is proposed to determine the risk profile of the population with the multi-step transmission, incorporating serial and parallel transmission pathways in the network. We address the strategies for identifying individuals for COVID-19 tests, based on risk assessment in the community. On testing a group of individuals, few of them can be positive, and the remaining can be negative, generating a set of possible test outcomes. The group for testing is identified with maximal variation in the risk profile, based on all possible test outputs. Further, the individuals at the risk are identified for different test outcomes that could be useful for alerting the right individuals, without their individual tests. Using the proposed testing framework, the strategist shall obtain much more prediction information, with a minimal number of tests. On systematic applicability, the present strategy can break the transmission chain, which in turn, can control the epidemic despite limited testing capacity.

## Data Availability

Algorithm of the code is provided.

## Acknowledgement

The author acknowledges Prof. Fabien Alibart and Prof. Giridhar U. Kulkarni for the encouragement.

## About the Author

Ankush Kumar received the M.S.-Ph.D. degree from the Jawaharlal Nehru Centre for Advanced Scientific Research, Bengaluru, in 2018, under the supervision of Prof. G. U. Kulkarni. He completed his first Post-Doctoral at Department of Mathematics, University of Pittsburgh and currently a Post-Doctoral Associate at Institut d’Électronique de Microélectronique et de Nanotechnologie, France with Prof. Fabien Alibart. His research interests are on modeling aspects of networks, devices, and neuromorphic systems.

## Notes

### Competing Interest Statement

The authors have declared no competing interest.

### Clinical Trial

Not applicable

### Funding Statement

Not applicable.

